# Prevalence of depression in elderly patients following acute coronary syndrome at discharge from Thong Nhat Hospital of Ho Chi Minh City, Vietnam

**DOI:** 10.1101/2025.05.06.25327091

**Authors:** Ngoc Vo, Cong Duc Nguyen, Thach Nguyen, Minh Huu Nhat Le

**Author notes:** **Corresponding Author:** Cong Duc Nguyen, Affiliation: Department of Geriatrics, Pham Ngoc Thach University of Medicine, Ho Chi Minh City, Vietnam., And Minh Huu Nhat Le, Affiliation: Interventional Cardiology, Methodist Hospital, Merrillville, IN, USA; St Mary Medical Center, Hobart, IN USA; Tan Tao University, School of Medicine, Long An Province, Vietnam.

## Abstract

**Background:** Depression is a common mental health issue among patients recovering from acute coronary syndrome (ACS), particularly often underdiagnosed in the older population. Early detection of depression at the time of discharge is crucial to minimize the risk of adverse outcomes.

**Objectives:** The primary aim of our study was to estimate the frequency of depression among older adults with acute coronary syndrome and identify relevant factors that are associated with depression at the time of discharge.

**Method of Study:** We conducted a descriptive cross-sectional study on 117 elderly patients with ACS discharged from Thong Nhat Hospital between March 2024 and June 2024. A Vietnamese version of the Geriatric Depression Scale 30-items (GDS–30) was administered to all participants to assess the depression prevalence. Statistical analyses were performed to describe sociodemographic and clinical characteristics related to depression. An estimate of the prevalence ratio (PR) and 95% confidence Intervals (95% CI) was obtained through multivariate Poisson regression with robust variance to determine the factors that are independently associated with depression.

**Results:** The prevalence of depression at discharge among elderly ACS patients was 15,4% (95% CI: 8,7% – 22,0%). In the multivariate regression analysis: female gender (PR = 4,36; 95% CI: 1,36 - 13,97), illiteracy (PR = 3,76; 95% CI: 1,61 - 8,76), high-risk CCI (PR = 3,6; 95% CI: 1,07 - 12,11), experiencing two or more stressful life events (PR = 3,69; 95% CI: 1,58 - 8,59), and low perceived social support (PR = 3,67; 95% CI: 1,73 – 7,82) were significantly associated with higher prevalence of depression.

**Conclusions:** Depression is a prevalent mental health issue among older adults following acute coronary syndrome events. This study highlights the prevalence of depressive symptoms in this population and emphasizes the importance of early detection and tailored intervention strategies for individuals predicted as being at high risk. Further research is warranted to explore the long-term consequences of post-ACS depression and to improve clinical outcomes and quality of life for elderly patients.

## INTRODUCTION

Vietnam is currently one of the fastest-growing aging populations in the world and is predicted to enter the aging population period by 2035. During 2019 and 2021, the proportion of elderly individuals (aged 60 years and older) rapidly increased from 11,86% to 12,80% of the total population.^1^ Although the average lifespan of Vietnamese people has increased, it has also led to a greater burden of diseases among the elderly, with depression being a prevalent mental health condition, ranking eighth among chronic illnesses in individuals aged 65 and older.^2^ Depression is a mood disorder characterized by persistent sadness, loss of interest or pleasure, sleep and appetite disturbance, psychomotor agitation or retardation, and thoughts of self-harm.^3^ Depression is often underrecognized in older adults due to differing symptoms, a reluctance to seek help, and the complex nature of their health conditions, which can lead to confusion with other physical illnesses.

Acute coronary syndrome (ACS) is a severe medical emergency that includes three clinical forms: ST-elevation myocardial infarction (STEMI), non-ST-elevation myocardial infarction (NSTEMI), and unstable angina (UA). It has been documented that after an ACS event, patients often experience psychological stress, with depression and anxiety being the most prevalent conditions.^4^ The bidirectional relationship between ACS and depression has been extensively studied. Approximately 20% of patients hospitalized for ACS meet the diagnostic criteria for depression according to *The Diagnostic and Statistical Manual of Mental Disorders, Fifth Edition* (DSM–5), and those who have experienced ACS events are 2 to 3 times more likely to develop mental health.^4,5^ Conversely, post-ACS depression is an independent risk factor for future cardiovascular events and mortality, primarily due to the reduction in treatment adherence.^6,7^

Given the limited data on the depression status among post-ACS elderly patients in Vietnam, this study aimed to investigate the prevalence of depression in this population and analyze its associations with sociodemographic factors and clinical features. By examining these relationships, we want to provide evidence-based insights to facilitate early detection and intervention for depression at the time of discharge. Ultimately, the findings aim to improve patient outcomes and enhance the quality of care for elderly individuals recovering from ACS.

## METHODS

### Study Design and Setting

This was a single-center, prospective cross-sectional study conducted in the Department of Emergency and Interventional Cardiology at Thong Nhat Hospital in Ho Chi Minh City, Vietnam between March 2024 and June 2024.

### Study Subjects

Patients were eligible for inclusion if they met the following criteria: (a) aged 60 years and older, (b) diagnosed with acute myocardial infarction (AMI) or unstable angina (UA), and (c) permitted for discharge by the physicians of the Department of Emergency and Interventional Cardiology at Thong Nhat Hospital. Exclusion criteria applied to patients who were unable to communicate effectively for interview completion, including those with impaired consciousness, dementia, or a history of psychiatric disorders that could affect the accuracy of information provided.

### Sample method

The sample size was estimated according to the following formula:

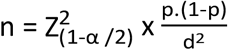

α: type 1 error probability, α= 0,05 → Z_(1-α/2)_ = 1,96;

d: tolerance, choose d = 0,08

p: the desired value of the ratio, we selected p = 26,4% according to a study by Nguyen Van Tan et al. (2021).^8^

A sample size of 117 patients was estimated to be adequate for detecting differences in primary outcomes, utilizing convenient sampling methods.

### Data collection

Data were collected using a structured research medical record. The questionnaire was administered through face-to-face interviews held one day before discharge by a single interviewer to minimize bias. Collected data included the patient sociodemographic information, clinical features and depressive condition.

### Variable definition

Depressive status was assessed using the Vietnamese version of GDS-30, which has been validated for high consistency, satisfactory reliability, and ease of understanding. It can be effectively used as a screening tool for depression in elderly patients in primary healthcare settings.^9^ The scale consists of 30 questions evaluating psychological experiences over the past two weeks, with responses scored as either “True” or “Not true”. The total GDS-30 score ranges from 0 to 30, and depression was classified into three levels: none to minimal (score: 0-9), mild (score: 10-19) and severe (score: 20-30). We selected a cut-off score of ≥ 10 as the optimal point for maximizing sensitivity without compromising specificity.^10,11^ The dependent variable was depression at discharge.

The severity of comorbidity was assessed using the Charlson comorbidity index (CCI), with a total score ≥ 3 indicating a high risk of one-year mortality.^12^ Functional impairment in activities of daily living (ADL) was evaluated using the Katz Activities of Daily Living (ADLs) scale. A score of ≤ 4 points on this scale denotes significant ADL impairment.^13^ Sleep disturbances were identified using the Pittsburgh Sleep Quality Index (PSQI), with a score of ≥ 5 indicating poor sleep quality.^14^ Psychological stress events were assessed based on the occurrence of stressful events that happened within the past 12 months, as well as significant life events experienced throughout the participant’s lifespan. A participant was considered to be affected if they reported experiencing two or more events from either of the scales.^15^ Perceived social support was measured with the Multidimensional Scale of Perceived Social Support (MSPSS). An average score below 5.1 on the MSPSS is considered indicative of low perceived social support from family, friends, and significant others.^16^

### Statistical Analyses

Data were entered into Microsoft Excel and exported to the SPSS version 27.0 statistical package for analysis. Descriptive statistics were reported as frequencies, percentages for categorical variables, and means with standard deviations (SD) for quantitative variables. Multivariate analyses using the Generalized Linear Model (GLM) were conducted to identify the determinants of depression. According to previous studies, the rate of post-ACS depression is expected to exceed 10%.^17,18^ In this context, reporting the odds ratio obtained from binary logistic regression in a cross-sectional study may lead to a significant overestimation of the strength of the relationship. As our response variable is binary, modified (Robust) Poisson regression was performed to obtain a more accurate estimate of the prevalence ratio, which is more appropriate for cross-sectional studies with binary outcomes.^19^ Initially, a univariate modified Poisson regression model was performed to identify variables that were statistically significantly associated with depression. These variables were subsequently included in a multivariable Poisson regression with robust standard errors to ascertain independent risk factors for post-ACS depression at discharge. Finally, the adjusted prevalence ratio (PR) and 95% confidence interval (CI) were estimated to illustrate the strength of the association. Variables with p-value < 0,05 were considered statistically significant factors.

### Ethical Considerations

The study was approved by the Ethics Committee for Medical and Biological Research of Pham Ngoc Thach University in Ho Chi Minh, Vietnam (Registration Number 1064/TDHYKPNT-HDDD). Approval date of IRB on March 6, 2024. Research period from March 15, 2024 to June 30, 2024.

## RESULTS

### Baseline characteristics of the patient population

A total of 117 patients with acute coronary syndrome completed the interview at the time of discharge. The study population had an average of 71,2 ± 7,4 years (age range 60-94), with the majority falling into the young-old age group (45,3%) and a higher proportion of males (59,8%). Most patients came from urban areas (64,1%), had religion (53%), currently lived with family members (94,4%), lived with a spouse (70,9%) and were no longer employed (67,5%). In addition, the widowhood rate was relatively high at 22,2%, while the illiteracy rate was low, accounting for 7,7%.

The majority of hospitalized patients were diagnosed with NSTEMI (55,6%), and 73,5% performed coronary angiography and/or intervention treatment during this admission. Most patients had no complications related to ACS (70,1%) and post-MI heart failure accounted for a significantly higher proportion (21,4%) among the recorded complications. Hypertension was the most prevalent chronic condition (80,3%) and about 30,8% of patients were predicted to be at high risk of being affected by multimorbidity based on the CCI. Limitations in ADLs (12%) and urinary incontinence (11,1%) had relatively low prevalence, but sleep disorders (confirmed by a PSQI score ≥ 5) were observed in nearly half of the study population (47,9%). Furthermore, 43,6% of patients reported experiencing ≥ 2 psychologically stressful life events, and 66,7% perceived a high level of social support. All characteristics are summarized in Table 1.

**Table 1.**
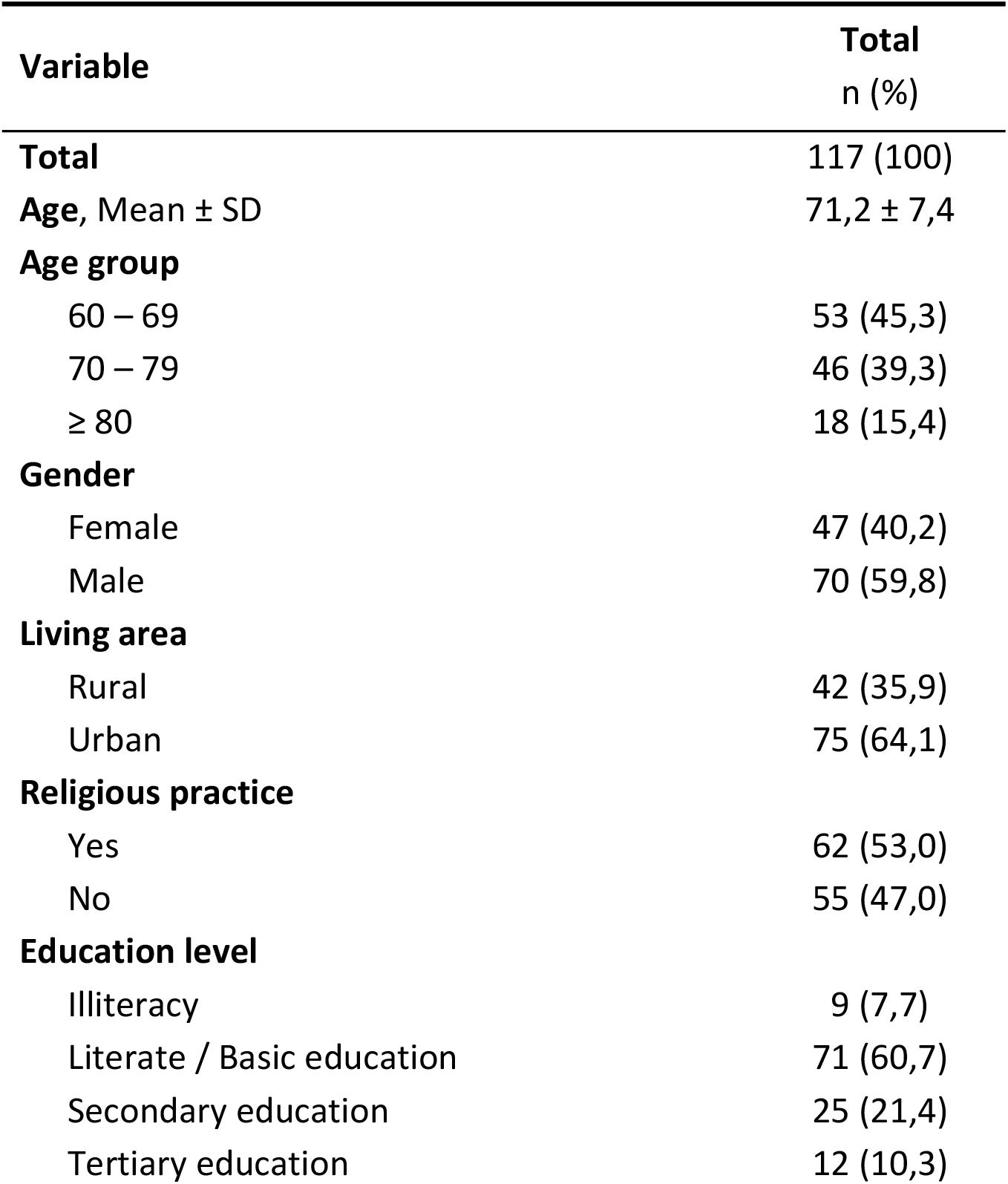
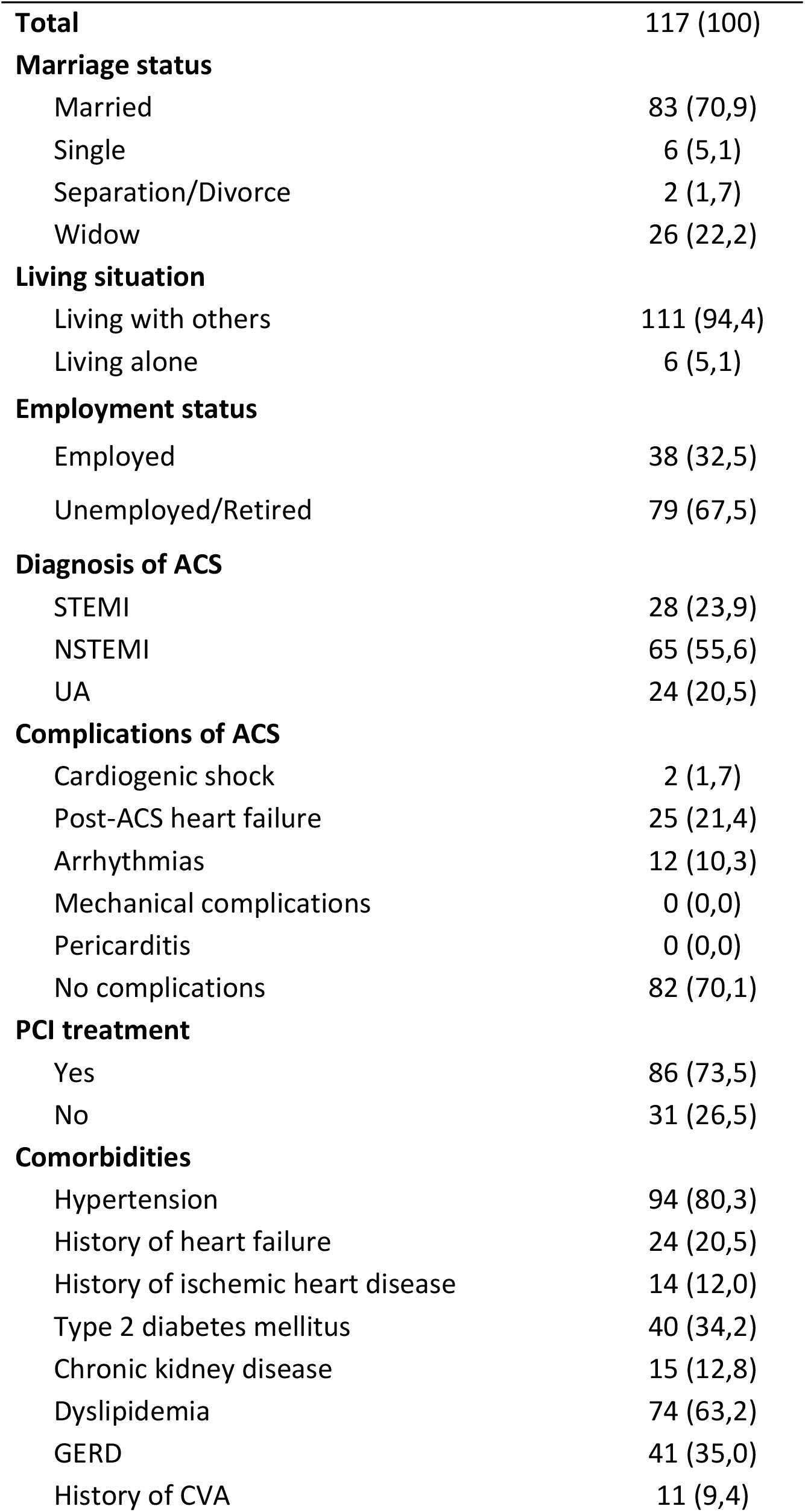
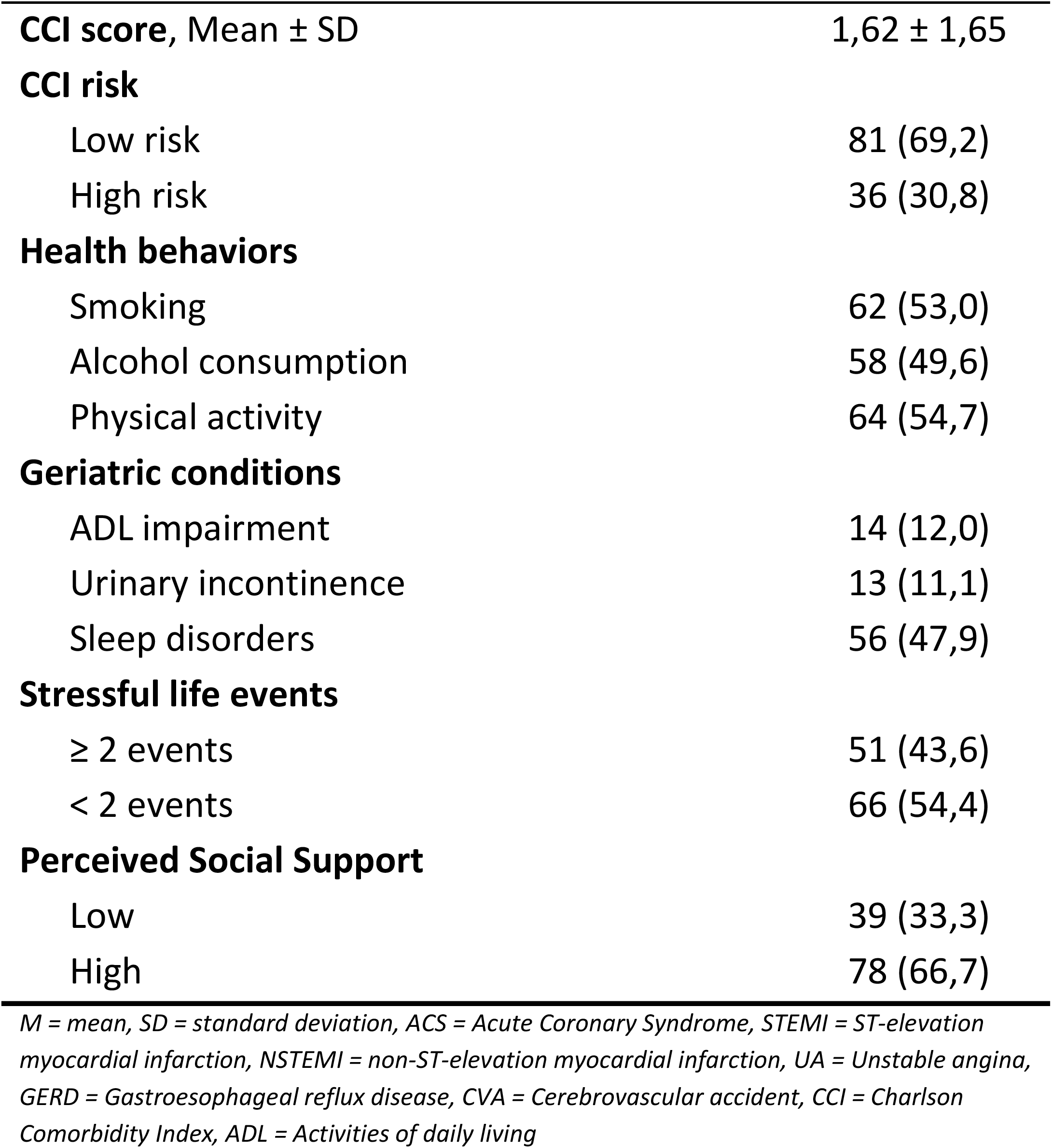
Sociodemographic and clinical characteristics of the study population.

| **Variable** | **Total**<br>**n (%)** |
| --- | --- |
| **Total** | 117 (100) |
| **Age, Mean ± SD** | 71,2 ± 7,4 |
| **Age group** |  |
| 60 – 69 | 53 (45,3) |
| 70 – 79 | 46 (39,3) |
| $\geq 80$ | 18 (15,4) |
| **Gender** |  |
| Female | 47 (40,2) |
| Male | 70 (59,8) |
| **Living area** |  |
| Rural | 42 (35,9) |
| Urban | 75 (64,1) |
| **Religious practice** |  |
| Yes | 62 (53,0) |
| No | 55 (47,0) |
| **Education level** |  |
| Illiteracy | 9 (7,7) |
| Literate / Basic education | 71 (60,7) |
| Secondary education | 25 (21,4) |
| Tertiary education | 12 (10,3) |
| **Total** | 117 (100) |
| **Marriage status** |  |
| Married | 83 (70,9) |
| Single | 6 (5,1) |
| Separation/Divorce | 2 (1,7) |
| Widow | 26 (22,2) |
| **Living situation** |  |
| Living with others | 111 (94,4) |
| Living alone | 6 (5,1) |
| **Employment status** |  |
| Employed | 38 (32,5) |
| Unemployed/Retired | 79 (67,5) |
| **Diagnosis of ACS** |  |
| STEMI | 28 (23,9) |
| NSTEMI | 65 (55,6) |
| UA | 24 (20,5) |
| **Complications of ACS** |  |
| Cardiogenic shock | 2 (1,7) |
| Post-ACS heart failure | 25 (21,4) |
| Arrhythmias | 12 (10,3) |
| Mechanical complications | 0 (0,0) |
| Pericarditis | 0 (0,0) |
| No complications | 82 (70,1) |
| **PCI treatment** |  |
| Yes | 86 (73,5) |
| No | 31 (26,5) |
| **Comorbidities** |  |
| Hypertension | 94 (80,3) |
| History of heart failure | 24 (20,5) |
| History of ischemic heart disease | 14 (12,0) |
| Type 2 diabetes mellitus | 40 (34,2) |
| Chronic kidney disease | 15 (12,8) |
| Dyslipidemia | 74 (63,2) |
| GERD | 41 (35,0) |
| History of CVA | 11 (9,4) |
| **CCI score, Mean ± SD** | 1,62 ± 1,65 |
| **CCI risk** |  |
| Low risk | 81 (69,2) |
| High risk | 36 (30,8) |
| **Health behaviors** |  |
| Smoking | 62 (53,0) |
| Alcohol consumption | 58 (49,6) |
| Physical activity | 64 (54,7) |
| **Geriatric conditions** |  |
| ADL impairment | 14 (12,0) |
| Urinary incontinence | 13 (11,1) |
| Sleep disorders | 56 (47,9) |
| **Stressful life events** |  |
| $\geq 2$ events | 51 (43,6) |
| $< 2$ events | 66 (54,4) |
| **Perceived Social Support** |  |
| Low | 39 (33,3) |
| High | 78 (66,7) |

*M = mean, SD = standard deviation, ACS = Acute Coronary Syndrome, STEMI = ST-elevation myocardial infarction, NSTEMI = non-ST-elevation myocardial infarction, UA = Unstable angina, GERD = Gastroesophageal reflux disease, CVA = Cerebrovascular accident, CCI = Charlson Comorbidity Index, ADL = Activities of daily living*

### Prevalence of Depression following ACS events

**Figure 1.**
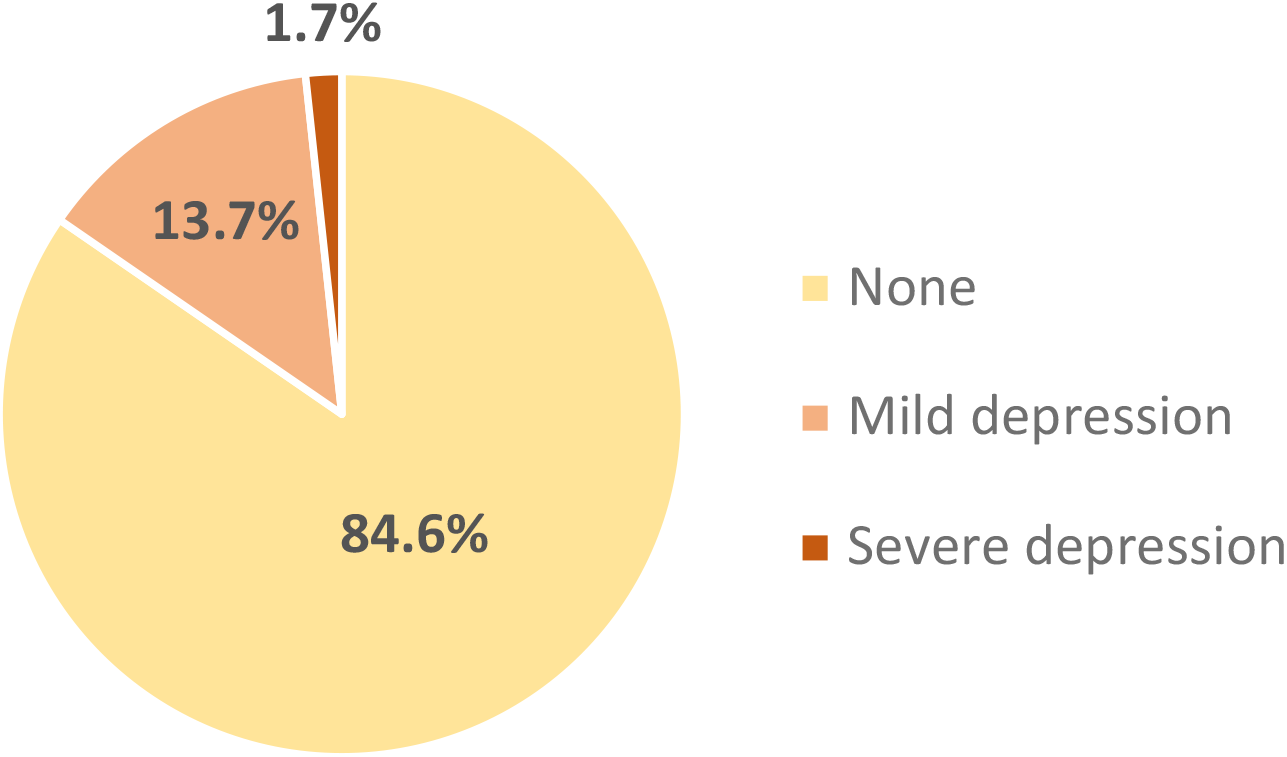
Prevalence of depression in post-ACS elderly patients at discharge

Based on the GDS-30 assessment, our study found a prevalence of 15,4% of elderly patients experiencing depression at the time of their discharge following ACS events, with 13,7% of them having mild depression (scores of 10 to 19), while only 1,7% were classified as severe depression (scores of 20 – 30). Separate analyses were performed to compare the different characteristics of elderly patients with depression to those without depression.

As shown in Table 2, the rate of depression was significantly higher among the female patients (p = 0,018), those who were illiterate (p = 0,006), and those who were currently living with their spouse (p = 0,036). Although we noted a greater prevalence of depression in patients aged 80 and older, as well as those living alone, these differences were not statistically significant.

**Table 2.** Univariate analysis of depression with sociodemographic characteristics of post-ACS elderly patients at discharge.

| Variable | Depression |  | Unadjusted PR<br>(95% CI) | p-value<sup>†</sup> |
| --- | --- | --- | --- | --- |
|  | Yes*<br>n (%) | No<br>n (%) |  |  |
| **Total** | 18 (100) | 99 (100) |  |  |
| **Age group** |  |  |  |  |
| 60 – 69<sup>‡</sup> | 5 (9,4) | 48 (90,6) | 1,00 | - |
| 70 – 79 | 8 (17,4) | 38 (82,6) | 1,84 (0,65 – 5,24) | 0,251 |
| ≥ 80 | 5 (27,8) | 18 (72,2) | 2,94 (0,96 – 9,01) | 0,058 |
| **Gender** |  |  |  |  |
| Female | 12 (25,5) | 35 (74,5) | 2,98 (1,20 – 7,38) | **0,018** |
| Male<sup>‡</sup> | 6 (8,6) | 64 (91,4) | 1,00 | - |
| **Living area** |  |  |  |  |
| Rural | 7 (16,7) | 35 (83,3) | 1,14 (0,48 – 2,71) | 0,773 |
| Urban<sup>‡</sup> | 11 (14,7) | 64 (85,3) | 1,00 | - |
| **Religious practice** |  |  |  |  |
| Yes | 10 (16,1) | 52 (83,9) | 1,11 (0,47 – 2,61) | 0,813 |
| No<sup>‡</sup> | 8 (14,5) | 47 (85,5) | 1,00 | - |
| **Illiteracy** |  |  |  |  |
| Yes | 4 (44,4) | 5 (55,6) | 3,43 (1,42 – 8,26) | **0,006** |
| No<sup>‡</sup> | 14 (13,0) | 94 (87,0) | 1,00 | - |
| **Married status** |  |  |  |  |
| Others<sup>a</sup> | 9 (26,5) | 25 (73,5) | 2,44 (1,06 – 5,62) | **0,036** |
| Married<sup>‡</sup> | 9 (10,8) | 74 (89,2) | 1,00 | - |
| **Living situation** |  |  |  |  |
| Living alone | 2 (33,3) | 4 (66,7) | 2,31 (0,68 – 7,83) | 0,178 |
| Living with others<sup>‡</sup> | 16 (14,4) | 95 (85,6) | 1,00 | - |
| **Employment status** |  |  |  |  |
| Unemployed/Retired | 14 (17,7) | 65 (82,3) | 1,68 (0,59 – 4,77) | 0,327 |
| Employed<sup>‡</sup> | 4 (10,5) | 34 (89,5) | 1,00 | - |

PR = Prevalence ratio, CI = Confidence Interval
\*: Based on Geriatric Depression Scale 30 items (GDS–30) ≥ 10.
<sup>†</sup>: Univariate Poisson regression with robust variance
<sup>‡</sup>: reference variable
<sup>a</sup>: including single, divorced, separate and widow status

**Table 3.**
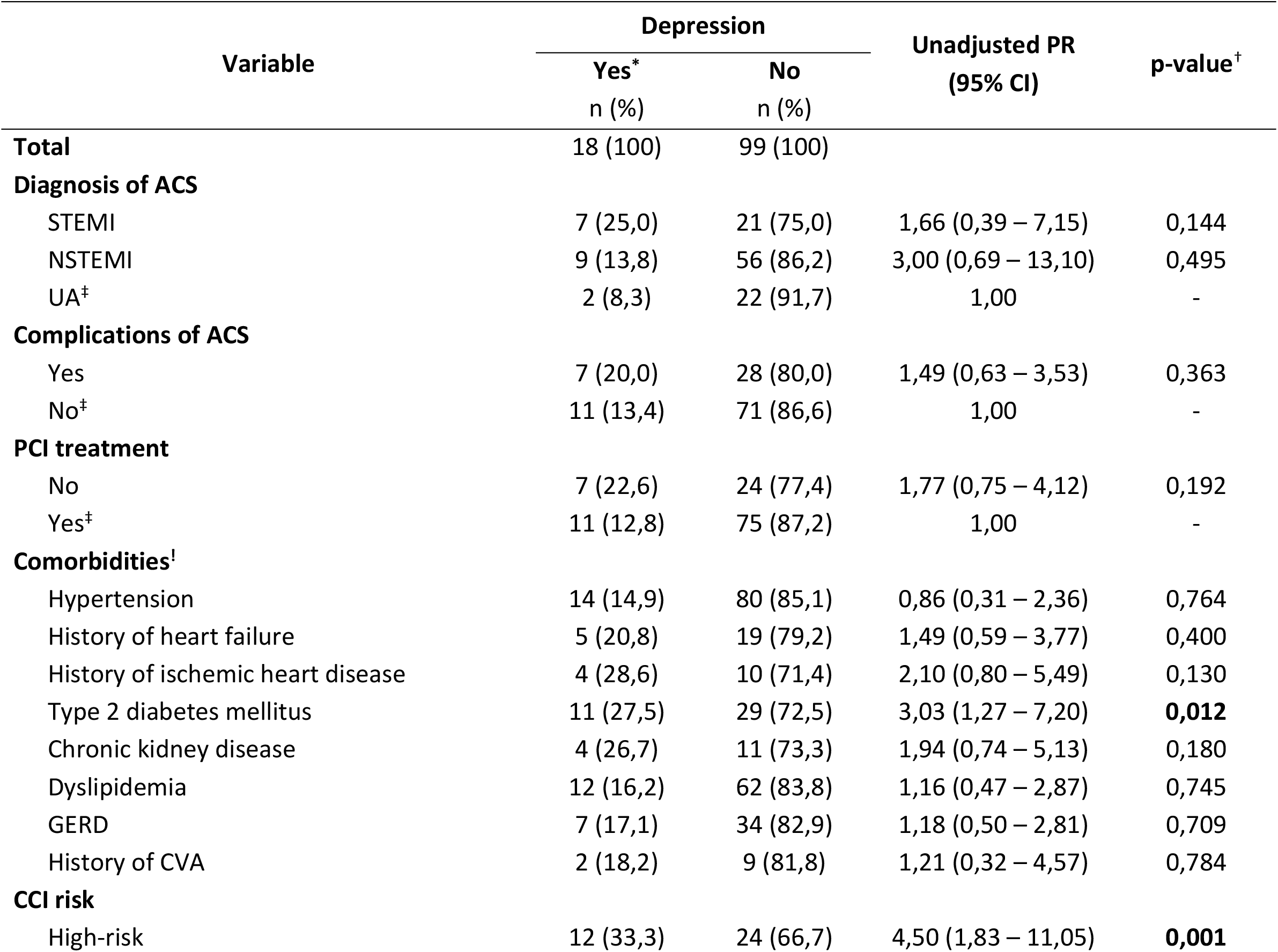
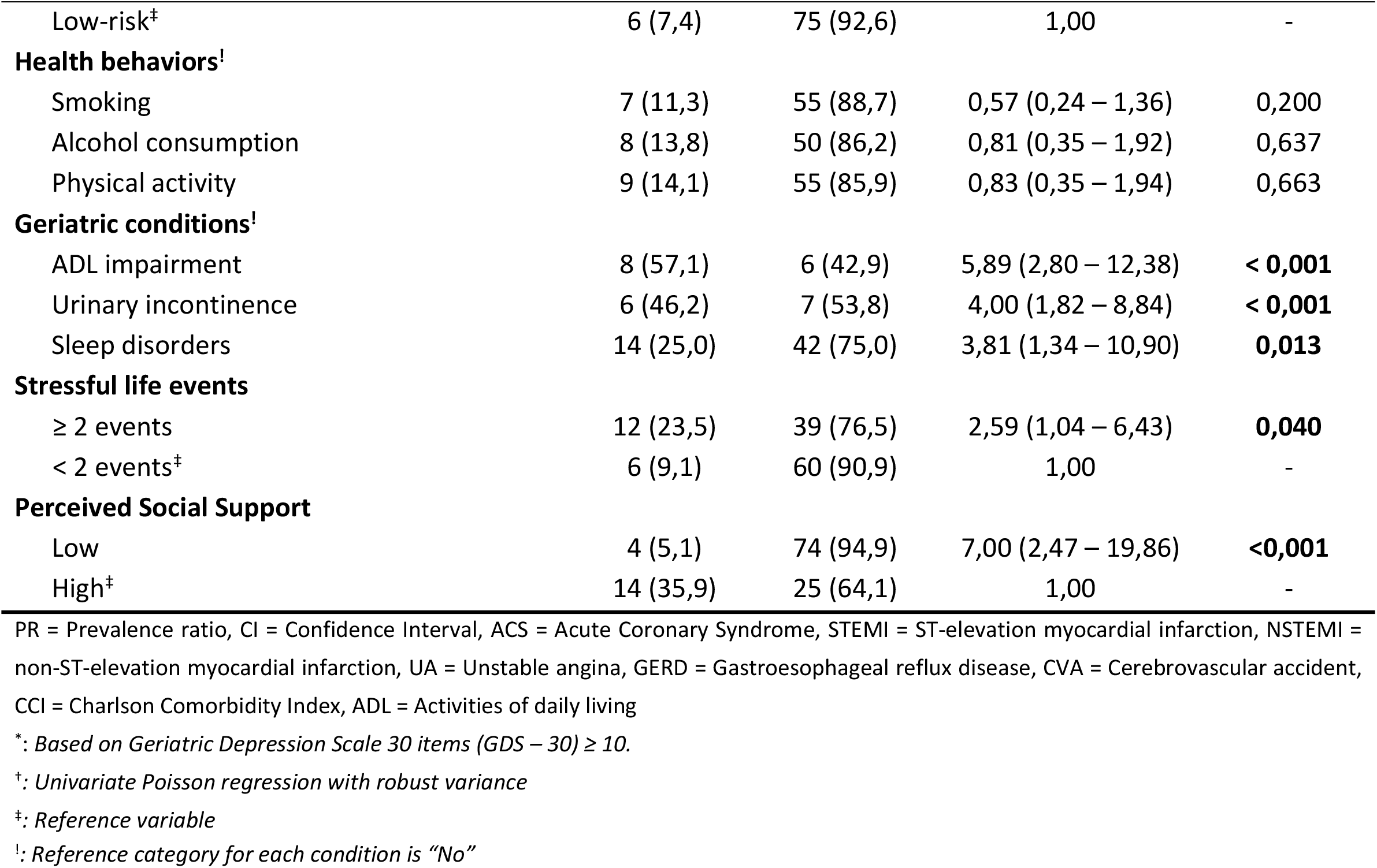
Univariate analysis of depression with clinical characteristics of post-ACS elderly patients at discharge.

**Table 4.**
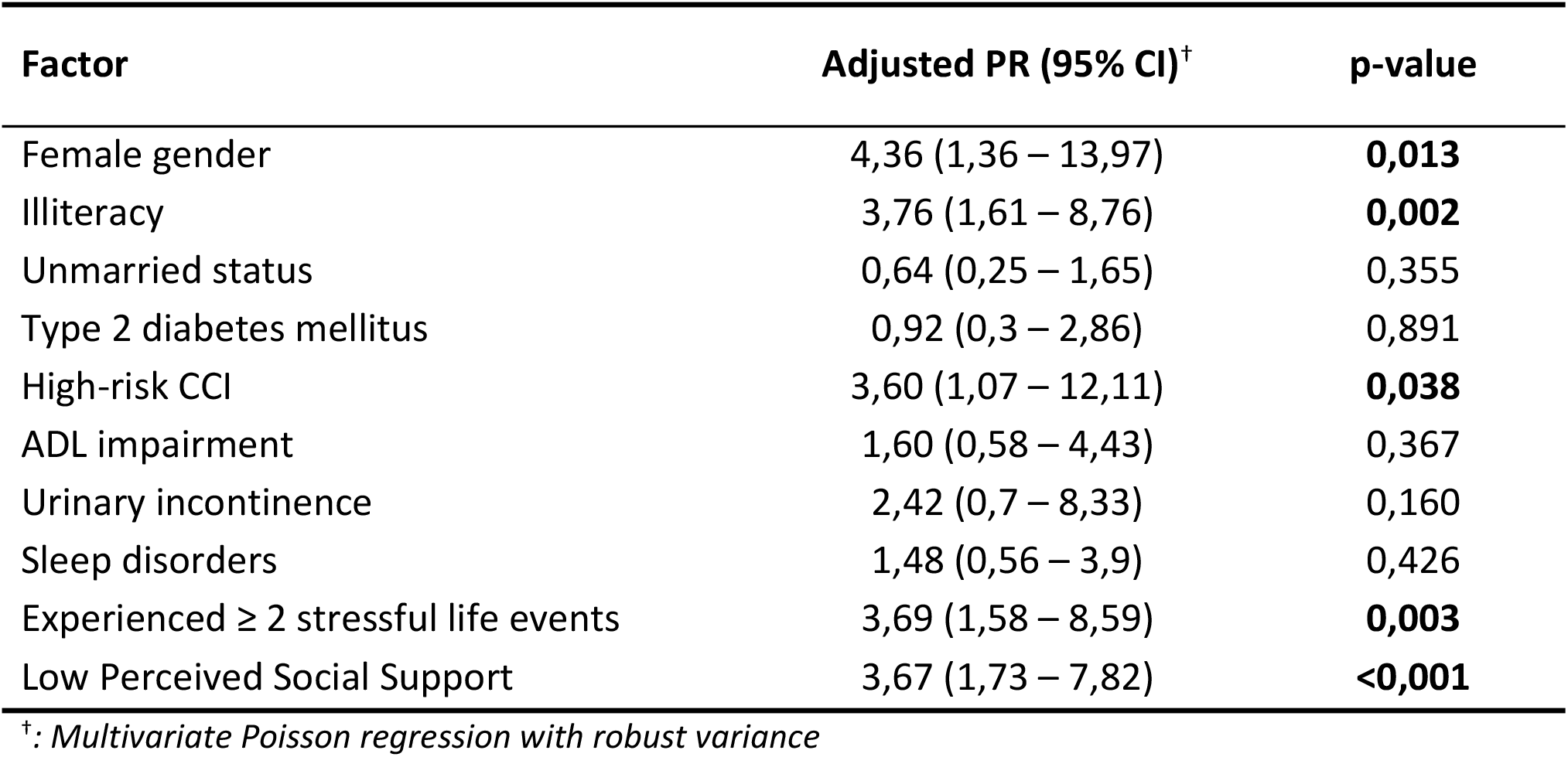
Multivariate modified Poisson regression of predictors of depression in post-ACS elderly patients at discharge.

Analyses of patient clinical characteristics show significant associations between depression and certain conditions. Depression was more likely among patients with type 2 diabetes (p = 0,012) and those considered at high risk of future mortality due to multimorbidity (confirmed by CCI score ≥ 3). Depression was also significantly more common in patients who had ADL impairment (p < 0,001), urinary incontinence issues (p < 0,001), and sleep disturbances (p = 0,013). Patients who had experienced more than one psychologically stressful event (either recent events within the past 12 months or major life events) were also more likely to have depression (p = 0,040). In contrast, patients with higher perceived social support (with average MSPSS scores ranging from 5,1 to 7) had a significantly lower prevalence of depression.

### Factors Associated with Outcomes

We selected 10 statistically significant variables for the multivariate regression model. After controlling for the influence of other covariates, we identified the following factors as significantly associated with post-ACS depression: female gender (PR = 4,36; 95% CI: 1,36 - 13,97), illiteracy (PR = 3,76; 95% CI: 1,61 - 8,76), high-risk CCI (PR = 3,6; 95% CI: 1,07 - 12,11), experiencing ≥ 2 stressful life events (PR = 3,69; 95% CI: 1,58 - 8,59), and low perceived social support (PR = 3,67; 95% CI: 1,73 – 7,82).

## DISCUSSION

### Prevalence of Depression

The study found that the prevalence of depression at the time of discharge among elderly patients recovering from ACS was 15,4% (95% CI: 8,7% – 22,0%) as measured by the GDS-30. A systematic review by Dong et al. (2024), derived from 28 studies, reported a pooled depression rate of 28,5% among ACS patients.^17^ A study by Thombs et al. (2003) indicated a post-AMI depression rate of 19,8%, which varied between 7,3% and 31,1% depending on the measurement tool used.^18^ When analyzing studies with similar elderly populations, we observed a gradual decline in depression rates over time. Specifically, Hayajneh et al. (2021) reported that 65,7% of elderly patients with AMI exhibited depressive symptoms at the time of emergency admission.^20^ In contrast, Romanelli (2002) and Nguyen Van Tan (2021) found depression rates of 22,9% and 26,4%, respectively.^8,21^ Our study revealed a significantly lower rate of depression at discharge, which may be attributed to patients feeling more reassured about their medical condition. This could explain the diminished reporting of certain items on the GDS-30 scale, such as feelings of sadness or fear of death. However, additional research is warranted to substantiate these psychological improvements relating to the admission phase. Notably, a considerable proportion of elderly patients exhibited signs of depression following acute coronary syndrome, underscoring the necessity for timely screening and intervention strategies.

### Factors Associated with Depression in Elderly Patients Post-AMI

Female patients exhibited a statistically significant depression rate that was 2,98 times higher than that of male patients. While meta-analyses indicate that the relationship between gender and depression in the elderly is not entirely consistent, many studies still report a higher incidence of depression among older women.^22,23^ Acciai (2017) recorded depression rates of 37,6% in men and 20,1% in women (p < 0,001).^24^ Similarly, Mallik S (2006) found that among patients aged 60 and older with AMI, depression was significantly more prevalent in women (21% vs. 15%, p = 0,02).^25^ Multivariate analysis in this study revealed that the female gender was independently associated with a 4,37 times higher prevalence of depression than the male group (95% CI: 1,36 – 13,97; p = 0,013). It is believed that older females may have a higher risk of depression, possibly due to hormonal imbalances after menopause and increased psychological stress from family and societal responsibilities, which may contribute to a greater likelihood of depression following ACS.

In our study, depression was more common among individuals with lower educational attainment, particularly among illiterate patients, who had a prevalence rate of 3,76 times higher (95% CI: 1,61 – 8,76; p = 0,006) than other groups. Illiteracy may create barriers to social communication, leading to feelings of inferiority and difficulty integrating into daily life.^25^ Additionally, patients without spouses reported a statistically higher prevalence of depression (PR = 2,44; p = 0,036). However, marital status lost its significance after controlling for other covariates in the multivariate regression model. This finding aligns with numerous surveys on depression in older adults.^8,22^ In contrast, religious factors, living area, living alone, and current occupational status did not demonstrate statistically significant relationships with depression in our study.

Type 2 Diabetes Mellitus was the only chronic condition showing a statistically significant association with depression (PR = 3,03; 95% CI: 1,27 – 7,20). Type 2 Diabetes Mellitus has been established as an independent risk factor for coronary artery disease and is closely related to depressive disorders, with depression rates among Type 2 Diabetes Mellitus patients reaching 40,2% to 48,2%.^26^ Furthermore, we found no association between a history of heart failure and depression, which differs from Nguyen Van Tan’s findings.^8^ This discrepancy may be attributed to population characteristics and the timing of the research.

Chronic multimorbidity has a strong correlation with depression, which is consistent with previous studies. Patients with ≥2 chronic conditions are 1,59 times more likely to have depression compared to those without any chronic diseases (adjusted OR = 1,595; 95% CI: 1,01 – 2,52; p = 0,045).^27^ Another study found a 32,5% depression rate among elderly individuals with ≥2 chronic conditions, compared to 8,2% in those with fewer conditions, with a 2,9-fold higher risk of depression (OR = 2,9; p < 0,001). Similar studies report depression risk ranging from 1,6 to 10,2 times higher in individuals with multimorbidity.^28^ In our study, high-risk patients had significantly higher depression rates than low-risk individuals (33,3% vs. 7,4%, PR = 4,50; p < 0,001). Multivariate analyses confirmed that multimorbidity is an independently associated factor, with a 3,6-fold increased risk of depression in the high-risk group (95% CI: 1,07 – 12,11; p = 0,038). Chronic health conditions may lead to dissatisfaction with quality of life, contributing to psychological stress and increasing the risk of depression.

Although geriatric conditions such as ADL impairment, urinary incontinence and sleep disturbances were noted to be statistically associated with depression, the multivariate analyses suggest that after controlling for other factors, these factors are no longer statistically significant in predicting depression. These findings align with the study by Nguyen Van Tan et al. (2021), which reported significant associations with ADL impairment and urinary incontinence but found that these conditions did not remain risk factors after conducting multivariate logistic regression.^8^ These issues may be common among the entire elderly population or could arise due to the influence of many underlying pathological factors that were not considered in the study, rather than simply being a symptom of depression.

Psychological factors have a significant impact on the prevalence of depression. Patients who reported experiencing two or more stressful life events were 3,69 times (95% CI: 1,58 – 8,59; p = 0,003) more likely to suffer from depression. According to Do Van Dieu et al. (2018), individuals who have experienced at least one “major life event in the past 12 months” or at least one “major life event in their lifetime” are independent risk factors that lead to a higher likelihood of developing depression, with adjusted odds ratios (OR) of 1,9 (p < 0,001) and 2,4 (p < 0,001), respectively. Several studies also report similar results, with odds ratios ranging from 1,6 to 3,3 (p < 0,05).^28^ Additionally, those identified as having low perceived social support are 3,67 times more likely to have depression (95% CI: 1,73 – 7,82; p < 0,001). This is understandable since social support is an important protective factor in maintaining mental health among elderly patients after an ACS event.

## LIMITATION

The study primarily relied on the GDS-30 scores to evaluate depression status without validating these findings against DSM-5 criteria. This approach may have introduced variability in diagnosing depression, as the GDS-30 focuses on symptomatology rather than providing a clinical diagnosis. In addition, this was a cross-sectional study that only assesses depression at the time of discharge and does not track the progression of depression as measured by the GDS-30 scale. Therefore, it cannot provide information on predictive risk factors for health outcomes in patients with depression.

## CONCLUSION

This study highlights the significant prevalence of depression among elderly patients recovering from ACS, with a rate of 15,4% at the time of discharge. The findings suggest that patients who are female, have illiteracy, present with multimorbidity, particularly type 2 diabetes mellitus, experience stressful life events, and have low perceived social support are more likely to suffer from depression. Overall, the results underscore the importance of the importance for increased awareness and proactive strategies to mitigate depression in elderly patients post-ACS. Future research should further investigate these associations and explore targeted interventions to address the complex factors contributing to depression in this population.

## Data Availability

All relevant data are within the manuscript and its Supporting Information files.

## Abbreviation

ACS: Acute Coronary Syndrome
AMI: Acute Myocardial Infarction
CCI: Charlson Comorbidity Index
MSPSS: Multidimensional Scale of Perceived Social Support
NSTEMI: Non-ST-elevation Myocardial Infarction
PCI: Percutaneous Coronary Intervention
PSQI: Pittsburgh Sleep Quality Index
UA: Unstable Angina
STEMI: ST-elevation Myocardial Infarction

## ACKNOWLEDGEMENTS

We would like to express our sincere gratitude to all the patients and their families who participated in this study. Their cooperation greatly enhanced our understanding of the depression status in elderly patients. We are especially thankful to the medical and research staff at Pham Ngoc Thach University for their unwavering dedication and hard work, which made this study possible. Finally, we would like to acknowledge Professor Dr. Cong Duc Nguyen, for introducing the topic, providing guidance and offering crucial support in the completion of this study.

